# LDL/Apo B ratio and Lp (a) Each Predict Coronary Artery Disease in Type2 Diabetes Independent of ASCVD Risk Score: A Case-Cohort Study

**DOI:** 10.1101/2020.09.16.20195990

**Authors:** Soghra Rabizadeh, Armin Rajab, Jeffrey I. Mechanick, Fatemeh Moosaie, Yekta Rahimi, Manouchehr Nakhjavani, Alireza Esteghamati

## Abstract

**Aim:** To evaluate the predictive value of the LDL-C/ApoB ratio for coronary heart disease (CHD) in patients with type2 diabetes (T2D).

**Methods:** In this case-cohort study, (apo)lipoproteins and glycemic indices were measured in 1058 individuals with T2D from February 2002 to March 2019, with a median duration of follow up of 10 years.

**Results:** Of 1058 patients with T2D, coronary heart disease occurred in 242 patients. Increased waist circumference, waist-to-hip ratio, and hemoglobin A1c, low-density lipoprotein cholesterol (LDL-C)/Apolipoprotein B (ApoB) ratio, presence of hypertension and metabolic syndrome, and insulin and statin use were more prevalent among patients with CHD (P<0.001). Logistic regression analysis showed that an LDL-C/ApoB ratio equal or lower than 1.2, as well as a Lp(a) cutoff level more than 25.45 mg/dl could each predict CHD independent of ASCVD risk score [adjusted OR:1.841, CI:1.257 – 2.698, P<0.001 and adjusted OR: 1.433, CI:1.014 – 2.026, P=0.041) respectively] when adjusted for multiple confounders.The atherogenic index of plasma (AIP) did not predict CHD.

**Conclusion:** This study showed that LDL-C/ApoB ratio and Lp(a) each, but not the atherogenic index of plasma, may be considered as an indicator of CHD independent of ASCVD risk score in patients with T2D. This finding merits further clarification to optimize preventive strategies for CHD.

## 1. Introduction

About one-third of patients with coronary heart disease (CHD) have diabetes, which confers a higher mortality risk compared to those patients without diabetes. Unfortunately, primary prevention of cardiovascular disease (CVD) remains inadequate in patients with diabetes[1, 2]. Not surprisingly, these figures translate into a significant healthcare burden in Iran, greatly impacted by cardiovascular events in patients with diabetes[3-5] [6].

LDL cholesterol (LDL-C) is a major risk factor for the development of atherosclerosis. This atherogenic risk depends on the amount of LDL-C and also on the number and size of the LDL particles[7, 8]. Small, dense LDL (sd-LDL) particles are highly atherogenic due to lower binding affinity for the LDL receptor, greater penetration into the arterial wall, prolonged half-life, and greater sensitivity to oxidative stress [9] [10].

There is one apolipoprotein B (ApoB) molecule in each LDL particle. Thus, the level of apoB represents the plasma number of LDL particles. Consequently, LDL-particle size is indirectly estimated by the LDL-C/ApoB ratio. Methods that are used to measure LDL-particle diameter are density gradient ultracentrifugation, nuclear magnetic resonance spectroscopy, and non-denaturing gradient gel electrophoresis. However, these methods are complex and relatively costly[11].

An LDL-C/ApoB ratio of 1.2 corresponds to an LDL-C diameter of 25.5 nm. This cutoff can distinguish the LDL-C pattern A (large buoyant LDL) from pattern B (sd-LDL)[11, 12]. In fact, the Québec Cardiovascular Study revealed the incidence of CHD increased significantly in patients with LDL-particle sizes of 25.5 nm or smaller [13].

The purpose of this study was to investigate the LDL/ApoB ratio as an indicator of small dense LDL for the prediction of CHD in a cohort of patients with type2 diabetes in Iran.

## 2. Material and methods

### 2.1. Study population

The current retrospective case-cohort study consists of 1058 patients with type 2 diabetes (T2D) who attended the diabetes clinic of Vali-Asr Hospital affiliated within Tehran University of Medical Sciences in Tehran, Iran, from February 2002 to March 2019. The exclusion criteria were having a history of overt diabetes complications, renal insufficiency (Cr > 2mg/dl or eGFR<30 cc/min), auto-immune or viral liver disease or liver cirrhosis, hypothyroidism, familial hypercholesterolemia, epilepsy, history of malignancies, hemoglobinopathy, history of any major mental disorders, history of known CHD, estrogen use by women, as well as pregnancy. Apolipoproteins, lipids, and glycemic indices were measured and followed every four months from 2002 to 2019. The study group was divided into two sub-groups including 816 patients without CHD and 242 patients with incident CHD.

Written informed consent was obtained from all study participants. The ethics committee of Tehran University of Medical Sciences approved the study. The study complied with the principles of the Declaration of Helsinki.

### 2.2. Physical examinations

Physical examinations were performed by trained staff. Height was measured with an inflexible measurement tape with a precision of 0.1 cm [14], Weight was measured using a portable digital scale (Tefal PP1100, France) with a precision of 0.1 kg while patients were asked to wear light clothing. Body mass index (BMI) (kg/m^2^) was calculated. After ten minutes of seated rest, blood pressure was measured three times, each five minutes apart, using calibrated Omron M7digital sphygmomanometers (Hoofddorp, The Netherlands) with the appropriate size of cuffs which covered at least 80% of the arm. The second and third records were used to calculate the mean value for systolic and diastolic blood pressure (SBP and DBP, respectively). Waist circumference (WC-mid) and hip circumference were measured by using a non-stretchable measuring tape with the subject standing still in a relaxed position, on a flat surface. “WC-mid” was measured as the horizontal plane midway between the costal margins and the iliac crest [15] [14],and hip circumference was measured as the distance around the largest part of the hip. The Waist to Hip Ratio (WHR) was calculated by dividing waist circumference by hip circumference. Demographic information, smoking, and medication history were obtained during the interview.

### 2.3. Laboratory evaluations

Ten mL of blood sample was drawn from each participants at 12–14h after an overnight fast. The samples were centrifuged (1500 rmp, for 10 min at standard room temperature 21 °C)[14] and stored in the temperature of −70 °C for laboratory evaluation. Serum concentrations of Lp(a), apoB, and apoA1 were measured using immunoturbidimetry (Beckman Coulter Inc., FL). Inter- and intra-assay coefficients of variation ranged from 1% to 3%.

Urinary albumin concentrations were analyzed by an immunoturbidimetric commercial kit (Randox, Antrim, UK). Hemoglobin A1c (A1C) was measured by high-performance liquid chromatography (A1C, DS5 Pink kit; Drew, Marseille, France). Fasting plasma glucose (FPG) and two-hour postprandial (2hPP) glucose measurements were performed using enzymatic colorimetric methods by the glucose oxidase test. The estimated glomerular filtration rate (eGFR) was calculated employing the Chronic Kidney Disease Epidemiology Collaboration Equation. Serum lipids concentration including total cholesterols, triglycerides (TG), high-density lipoprotein cholesterol (HDL-C), and low-density lipoprotein cholesterol (LDL-C)[16], were measured using enzymatic methods[17]. The atherogenic index of plasma (AIP) was calculated as”Logarithm10 (TG/HDL-C)”[18], and non-HDL-C as total cholesterol minus HDL-C.

### 2.4. Assessment of ASCVD (Atherosclerotic cardiovascular disease) 10-year risk score and metabolic syndrome

The ASCVD 10 year risk score was calculated based on onMcClellandet al.[19] formula. Metabolic syndrome status was determined based on NCEP ATP III criteria (The US National Cholesterol Education Programme Adult Treatment Panel III). [20]Also, the Iran national recommendation for WC>90 cm both in males and females was used, according to the International Diabetes Federation criteria[21, 22].

### 2.5. Assessment of outcomes, CHD and microvascular complications

Coronary heart disease was defined as having a percutaneous coronary intervention (PCI), a coronary artery bypass graft (CABG), acute coronary syndrome, or myocardial infarction. Microvascular complications (e.g., diabetic retinopathy, neuropathy, and nephropathy)were evaluated according to ADA recommendations.[23]

### 2.6. Statistical Analysis

The Statistical Package for Social Science program(SPSS for Windows Inc. version 22. Chicago, Illinois, USA) was used for statistical analysis. Kolmogorov–Smirnov and Shapiro-Wilk normality tests, P-P plot, and histogram were used to test the normality of the study subjects. To assess the association between different variables, Uni-variable analysis of potential continuous and categorical variables was performed using t-test and Chi-square, respectively. Mean ± standard deviation(SD) are reported for continuous variables and categorical variables are presented as frequency and percentage (%). Unadjusted odds ratio and mean difference are presented for categorical and continuous variables, respectively. Binary logistic regression was performed on the variables with an unadjusted P-value<0.2 for CHD. The adjustment was done separately for confounding variables of CHD (i.e., age, gender, systolic and diastolic blood pressure, BMI, smoking status, statin use, duration of diabetes, type of diabetes medication [oral/insulin], A1C, triglyceride, waist circumference, WHR, eGFR, non-HDL-C/HDL-C ratio, and apoB/apoA1 ratio). The correlation matrix was used to predict the collinearity among predictors. Bivariate Pearson’s correlation analysis was performed to assess the correlations of AIP and other variables, and multinomial regression analysis was used to assess the relation of AIP and ASCVD 10-year risk score categories. The Receiver Operating Characteristics (ROC) curve analysis was performed and the Youden index was employed to find the predictive cut-off of Lp(a) for CHD. A P-value≤0.05 was regarded as statistically significant.

## 3. Results

### 3.1. Baseline characteristics

One thousand fifty-eight eligible subjects including 559 women and 499 men were followed in this study. During the follow-up period, CHD occurred in 242 patients in this cohort. The baseline characteristics of the participants are shown in Tables 1 and 2. Coronary heart disease occurred more frequently in men, older subjects, and those with a greater duration of diabetes (P<0.001). Also, patients with CHD had a higher prevalence of hypertension,increased waist circumference, WHR, elevated A1C, low LDL-C/Apo B ratio, and greater insulin and statin use than those without CHD (P<0.001). The prevalence of metabolic syndrome based on NCEP ATP III criteria was not significantly different regarding CHD status (P=0.081); however, according to the Iranian cutoff for WC (both in females and males> 90cm), the prevalence of metabolic syndrome was significantly higher among patients with CHD (82.6% vs. 76.1%, P=0.036). Moreover, patients with CHDhad significantly lower levels of total cholesterol, apo B lipoprotein, non-HDL-C, non-HDL-C/HDL-C ratio, LDL-C, triglyceride, and eGFR, and higher levels of Lp(a) and systolic blood pressure than individuals without CHD (P<0.05). Although the level of HDL-C was lower among patients with CHD(P<0.001), the prevalence of low HDL-C was not different between groups (P=0.255).Furthermore, there no significant differences in FBS, 2hpp, HOMA-IR, CRP, apo A1 lipoprotein, apoB/apoA1 ratio, AIP, diastolic blood pressure, and smoking status between groups (P>0.05). Additionally, at the end of follow-up, the prevalence of microvascular complications of diabetes was higher among patients with CHD (65.3% vs. 53.7%, P=0.001). At baseline, individuals who developed CHD had a higher calculated 10-year risk of CHD based on the ASCVD risk score (P<0.001). Also, the prevalence of diabetes microvascular complications during follow-up was higher among patients with χ CHD (OR=1.62, P= 0.001). Prevalence of low (<1.2) LDL-C/Apo B ratio was54.9% (n=307) among females vs 59.3% (n=296) among males (χ^2^ =1.006, P=0.159).

**Table 1.**
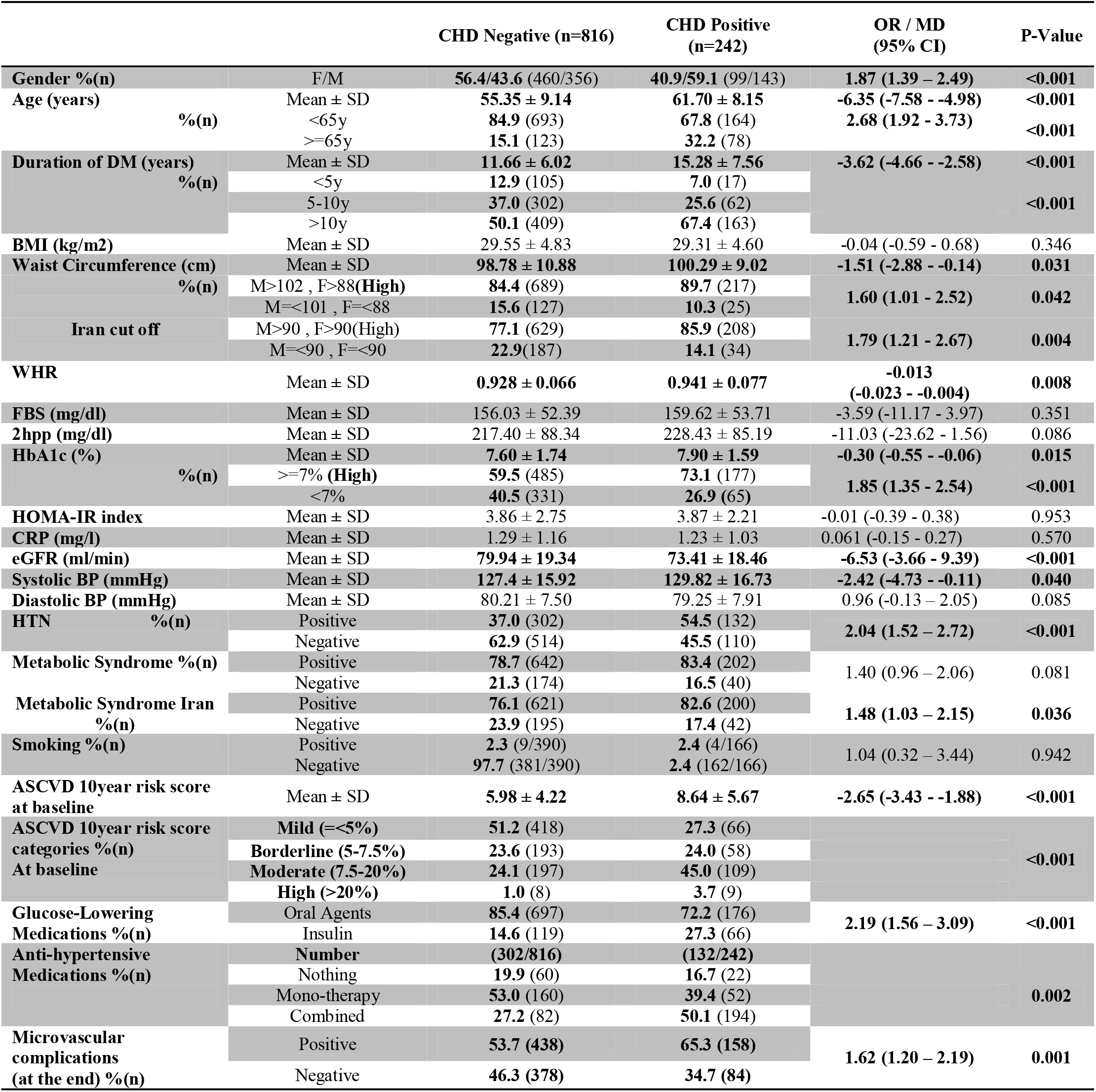
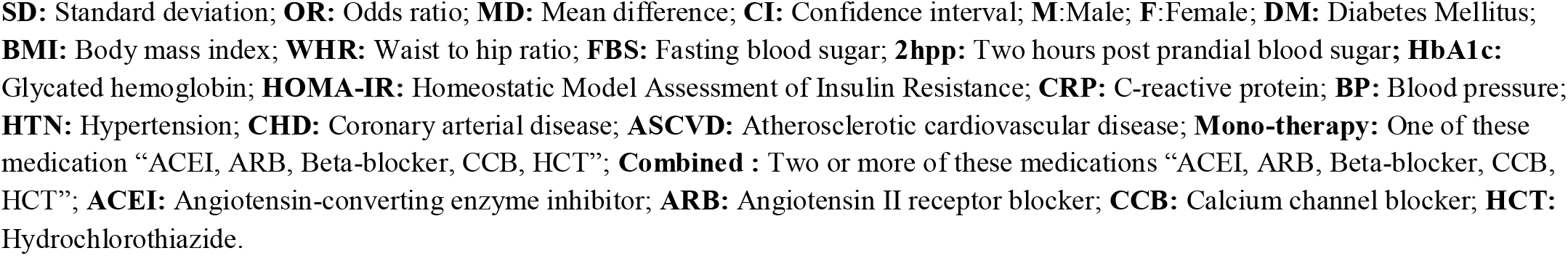
Baseline characteristics of study population except lipid profiles based on CHD status

|  |  | CHD Negative (n=816) | CHD Positive (n=242) | OR / MD (95% CI) | P-Value |
| --- | --- | --- | --- | --- | --- |
| **Gender % (n)** | F/M | **56.4/43.6** (460/356) | **40.9/59.1** (99/143) | **1.87 (1.39 – 2.49)** | **<0.001** |
| **Age (years)** | Mean ± SD | **55.35 ± 9.14** | **61.70 ± 8.15** | **-6.35 (-7.58 - -4.98)** | **<0.001** |
|  | % (n) |  |  |  |  |
|  | <65y | **84.9** (693) | **67.8** (164) | **2.68 (1.92 - 3.73)** | **<0.001** |
|  | ≥65y | **15.1** (123) | **32.2** (78) |  |  |
| **Duration of DM (years)** | Mean ± SD | **11.66 ± 6.02** | **15.28 ± 7.56** | **-3.62 (-4.66 - -2.58)** | **<0.001** |
|  | % (n) |  |  |  |  |
|  | <5y | **12.9** (105) | **7.0** (17) |  |  |
|  | 5-10y | **37.0** (302) | **25.6** (62) |  | **<0.001** |
|  | >10y | **50.1** (409) | **67.4** (163) |  |  |
| **BMI (kg/m<sup>2</sup>)** | Mean ± SD | 29.55 ± 4.83 | 29.31 ± 4.60 | -0.04 (-0.59 - 0.68) | 0.346 |
| **Waist Circumference (cm)** | Mean ± SD | **98.78 ± 10.88** | **100.29 ± 9.02** | **-1.51 (-2.88 - -0.14)** | **0.031** |
|  | % (n) |  |  |  |  |
|  | M>102 , F>88(High) | **84.4** (689) | **89.7** (217) | **1.60 (1.01 - 2.52)** | **0.042** |
|  | M≤101 , F≤88 | **15.6** (127) | **10.3** (25) |  |  |
| **Iran cut off** | M>90 , F>90(High) | **77.1** (629) | **85.9** (208) | **1.79 (1.21 - 2.67)** | **0.004** |
|  | M≤90 , F≤90 | **22.9** (187) | **14.1** (34) |  |  |
| **WHR** | Mean ± SD | **0.928 ± 0.066** | **0.941 ± 0.077** | **-0.013 (-0.023 - -0.004)** | **0.008** |
| **FBS (mg/dl)** | Mean ± SD | 156.03 ± 52.39 | 159.62 ± 53.71 | -3.59 (-11.17 - 3.97) | 0.351 |
| **2hpp (mg/dl)** | Mean ± SD | 217.40 ± 88.34 | 228.43 ± 85.19 | -11.03 (-23.62 - 1.56) | 0.086 |
| **HbA1c (%)** | Mean ± SD | **7.60 ± 1.74** | **7.90 ± 1.59** | **-0.30 (-0.55 - -0.06)** | **0.015** |
|  | % (n) |  |  |  |  |
|  | ≥7% (High) | **59.5** (485) | **73.1** (177) | **1.85 (1.35 - 2.54)** | **<0.001** |
|  | <7% | **40.5** (331) | **26.9** (65) |  |  |
| **HOMA-IR index** | Mean ± SD | 3.86 ± 2.75 | 3.87 ± 2.21 | -0.01 (-0.39 - 0.38) | 0.953 |
| **CRP (mg/l)** | Mean ± SD | 1.29 ± 1.16 | 1.23 ± 1.03 | 0.061 (-0.15 - 0.27) | 0.570 |
| **eGFR (ml/min)** | Mean ± SD | **79.94 ± 19.34** | **73.41 ± 18.46** | **-6.53 (-3.66 - 9.39)** | **<0.001** |
| **Systolic BP (mmHg)** | Mean ± SD | **127.4 ± 15.92** | **129.82 ± 16.73** | **-2.42 (-4.73 - -0.11)** | **0.040** |
| **Diastolic BP (mmHg)** | Mean ± SD | 80.21 ± 7.50 | 79.25 ± 7.91 | 0.96 (-0.13 - 2.05) | 0.085 |
| **HTN % (n)** | Positive | **37.0** (302) | **54.5** (132) | **2.04 (1.52 – 2.72)** | **<0.001** |
|  | Negative | **62.9** (514) | **45.5** (110) |  |  |
| **Metabolic Syndrome % (n)** | Positive | **78.7** (642) | **83.4** (202) | 1.40 (0.96 – 2.06) | 0.081 |
|  | Negative | **21.3** (174) | **16.5** (40) |  |  |
| **Metabolic Syndrome Iran % (n)** | Positive | **76.1** (621) | **82.6** (200) | **1.48 (1.03 – 2.15)** | **0.036** |
|  | Negative | **23.9** (195) | **17.4** (42) |  |  |
| **Smoking % (n)** | Positive | **2.3** (9/390) | **2.4** (4/166) | 1.04 (0.32 – 3.44) | 0.942 |
|  | Negative | **97.7** (381/390) | **2.4** (162/166) |  |  |
| **ASCVD 10year risk score at baseline** | Mean ± SD | **5.98 ± 4.22** | **8.64 ± 5.67** | **-2.65 (-3.43 - -1.88)** | **<0.001** |
| **ASCVD 10year risk score categories % (n) At baseline** | Mild (≤5%) | **51.2** (418) | **27.3** (66) |  |  |
|  | Borderline (5-7.5%) | **23.6** (193) | **24.0** (58) |  | **<0.001** |
|  | Moderate (7.5-20%) | **24.1** (197) | **45.0** (109) |  |  |
|  | High (>20%) | **1.0** (8) | **3.7** (9) |  |  |
| **Glucose-Lowering Medications % (n)** | Oral Agents | **85.4** (697) | **72.2** (176) | **2.19 (1.56 – 3.09)** | **<0.001** |
|  | Insulin | **14.6** (119) | **27.3** (66) |  |  |
| **Anti-hypertensive Medications % (n)** | Number | **(302/816)** | **(132/242)** |  |  |
|  | Nothing | **19.9** (60) | **16.7** (22) |  |  |
|  | Mono-therapy | **53.0** (160) | **39.4** (52) |  | **0.002** |
|  | Combined | **27.2** (82) | **50.1** (194) |  |  |
| **Microvascular complications (at the end) % (n)** | Positive | **53.7** (438) | **65.3** (158) | **1.62 (1.20 – 2.19)** | **0.001** |
|  | Negative | **46.3** (378) | **34.7** (84) |  |  |

**SD:** Standard deviation; **OR:** Odds ratio; **MD:** Mean difference; **CI:** Confidence interval; **M:** Male; **F:** Female; **DM:** Diabetes Mellitus; **BMI:** Body mass index; **WHR:** Waist to hip ratio; **FBS:** Fasting blood sugar; **2hpp:** Two hours post prandial blood sugar; **HbA1c:** Glycated hemoglobin; **HOMA-IR:** Homeostatic Model Assessment of Insulin Resistance; **CRP:** C-reactive protein; **BP:** Blood pressure; **HTN:** Hypertension; **CHD:** Coronary arterial disease; **ASCVD:** Atherosclerotic cardiovascular disease; **Mono-therapy:** One of these medication “ACEI, ARB, Beta-blocker, CCB, HCT”; **Combined :** Two or more of these medications “ACEI, ARB, Beta-blocker, CCB, HCT”; **ACEI:** Angiotensin-converting enzyme inhibitor; **ARB:** Angiotensin II receptor blocker; **CCB:** Calcium channel blocker; **HCT:** Hydrochlorothiazide.

**Table 2.** Baseline lipid Parameters of study population based on CHD status

|  |  | CHD Negative<br>(n=816) | CHD Positive<br>(n=242) | OR / MD<br>(95% CI) | P-Value |
| --- | --- | --- | --- | --- | --- |
| **Gender % (n)** | F/M | **56.4/43.6** (460/356) | **40.9/59.1** (99/143) | **1.87 (1.39 – 2.49)** | **<0.001** |
| **Cholesterol (mg/dl)** | Mean ± SD | **189.65 ± 41.37** | **166.45 ± 38.19** | **23.19 (18.25 - 30.63)** | **<0.001** |
| **Apo B (g/l)** | Mean ± SD | **92.79 ± 26.34** | **85.97 ± 25.14** | **6.83 (3.06 - 10.59)** | **<0.001** |
| **ApoA1 (g/l)** | Mean ± SD | 131.99 ± 29.74 | 129.70 ± 25.83 | 2.29 (-1.93 - 6.52) | 0.286 |
| **Apo B/ApoA1 ratio** | Mean ± SD | 0.75 ± 0.34 | 0.71 ± 0.32 | 0.04 (-0.01 - 0.09) | 0.092 |
| **Lp(a) (mg/dl)** | Mean ± SD | **35.18 ± 29.78** | **46.46 ± 37.19** | **-11.28**<br>**(-16.45 - -6.12)** | **<0.001** |
| **HDL-C (mg/dl)** | Mean ± SD | **46.23 ± 11.85** | **41.65 ± 10.63** | **3.29 (1.43 - 5.00)** | **<0.001** |
| **% (n)** | M<40 , F<50 (**Low**) | 52.5 (428) | 56.6 (137) | 1.18 (0.89 - 1.58) | 0.255 |
|  | M>40 , F>50 | 47.5 (388) | 43.4 (105) |  |  |
| **Non HDL-C (mg/dl)** | Mean ± SD | **145.33 ± 42.89** | **125.18 ± 37.11** | **20.15 (14.02 – 26.27)** | **<0.001** |
| **Non HDL-C/HDL-C Ratio** | Mean ± SD | **3.32 ± 1.24** | **3.09 ± 1.23** | **0.23 (0.04 – 0.41)** | **0.015** |
| **LDL-C (mg/dl)** | Mean ± SD | **109.77 ± 35.15** | **91.67 ± 31.25** | **18.09 (13.17 – 23.02)** | **<0.001** |
| **LDL-C/ApoB Ratio** | Mean ± SD | **1.24 ± 0.46** | **1.13 ± 0.46** | **0.11 (0.05 – 0.18)** | **0.001** |
| **% (n)** | <1.2 (**Low**) | **54.0** (441) | **66.9** (162) | **1.73 (1.28 – 2.33)** | **<0.001** |
|  | ≥1.2 | **46.0** (375) | **33.0** (80) |  |  |
| **Triglyceride (mg/dl)** | Mean ± SD | **174.79 ± 87.51** | **160.54 ± 77.74** | **14.25 (1.93 – 26.57)** | **0.023** |
| **% (n)** | >150 (**High**) | **53.9** (440) | **46.7** (113) | **0.749 (0.56 - 0.99)** | **0.048** |
|  | <150 | **46.1** (376) | **53.3** (129) |  |  |
| **AIP** | Mean ± SD | 0.55 ± 0.26 | 0.54 ± 0.25 | 0.005 (-0.03 – 0.04) | 0.803 |
| **Statins use % (n)** | Nothing | **43.4** (354) | **30.6** (74) | **1.74 (1.28 – 2.36)** | **<0.001** |
|  | Use statins | **56.6** (462) | **69.4** (168) |  |  |

**SD:** Standard deviation; **OR:** Odds ratio; **MD:** Mean difference; **CI:** Confidence interval; **M:** Male; **F:** Female; **ApoB:** apolipoprotein B; **ApoA1:** apolipoprotein A1; **Lp(a):** Lipoprotein(a); **HDL-C:** High density lipoprotein cholesterol; **LDL-C:** Low density lipoprotein cholesterol; **AIP:** Atherogenic index of plasma.

### 3.2. LDL-C/Apo B ratio and CHD

The level of LDL-C/Apo B ratio was significantly lower among patients with CHD (P=0.001) (Figure 1). Also, the number of patients with a low LDL-C/Apo B ratio (≤1.2) was significantly greater among patients with CHD (P<0.001) (Figure 2). Logistic regression analysis was performed to determine the predictability of CHD based on the LDL-C/Apo B ratio and to eliminate the impacts of confounders (Table3). The analysis revealed thatonlyaLDL-C/Apo B ratio≤1.2 and elevated Lp(a) could each predict CHD independent of ASCVD score (respectively OR=1.841, P=0.002; OR=1.009, P<0.001; respectively) when adjusted for gender, age, systolic and diastolic blood pressure, A1C, smoking status, BMI, statin use, duration of diabetes, type of diabetes medication (oral/insulin), triglyceride, Waist circumference, WHR, eGFR, non-HDL-C/HDL-C ratio, and apoB/apoA1 ratio. Interaction analysis showed no significant interaction between the ASCVD 10year risk score and LDL-C/Apo B ratio (P=0.899).

**Figure 1.**
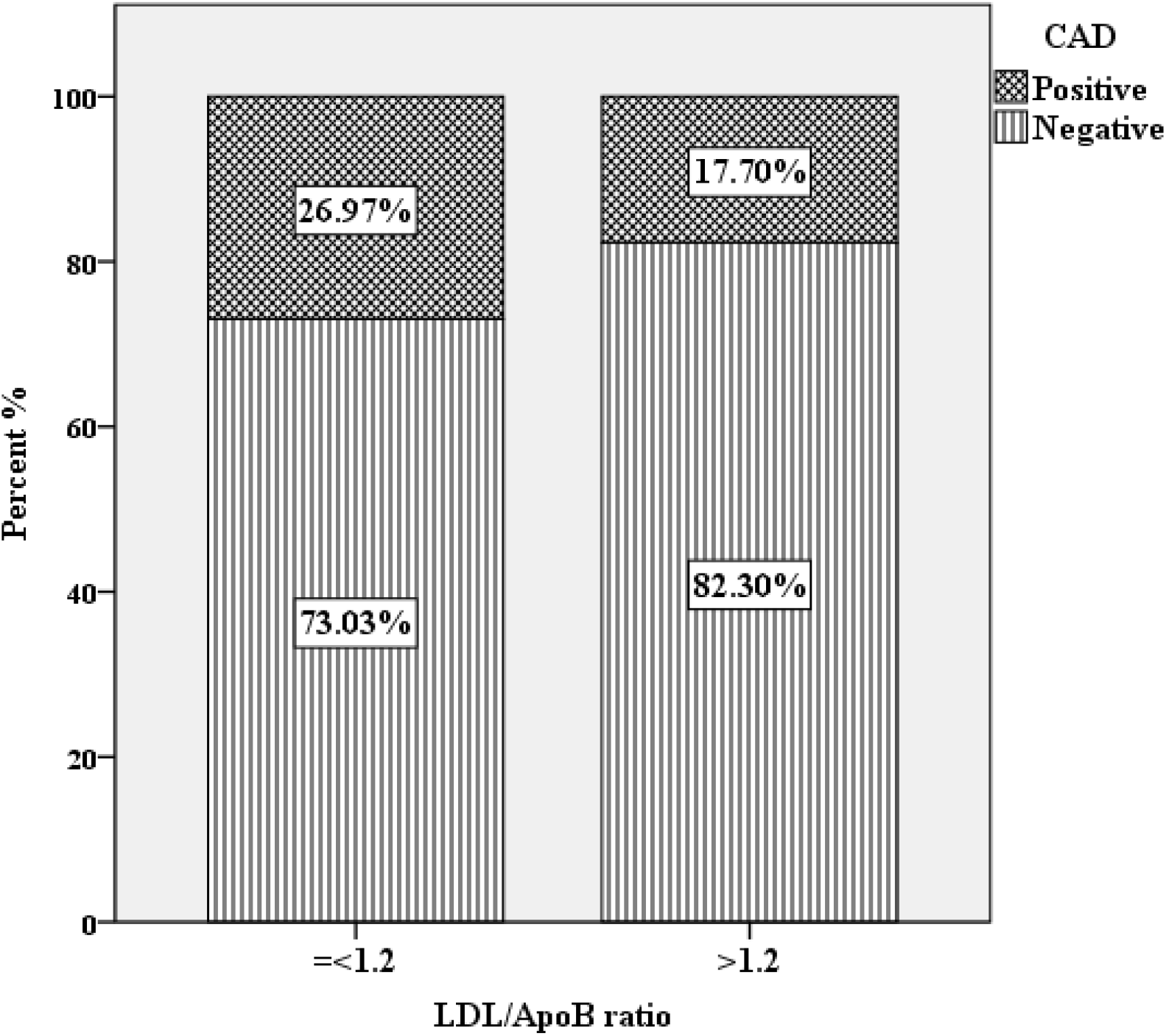
Comparison of the small sized LDL particles based on CHD status. **LDL/ApoB ratio:** Low density lipoprotein cholesterol **/** Apolipoprotein B**; CHD:** Coronary arterial disease

The ROC curve analysis showed that Lp(a) could predict the CHD (AUC=0.588, P<0.001), and the maximum Youden index for an optimal cutoff was Lp(a)=25.45 mg/dl, with a sensitivity=61.5%, andspecificity=50.0%. After the categorization of Lp(a), the new ORs were shown in table 3.The results of the analysis of the possibility of apoB predictive value for CHD in a separate regression model adjusted for possible confounders was not significant (OR=0.997; CI95%= 0.990-1.004; P=0.303).

**Table 3.**
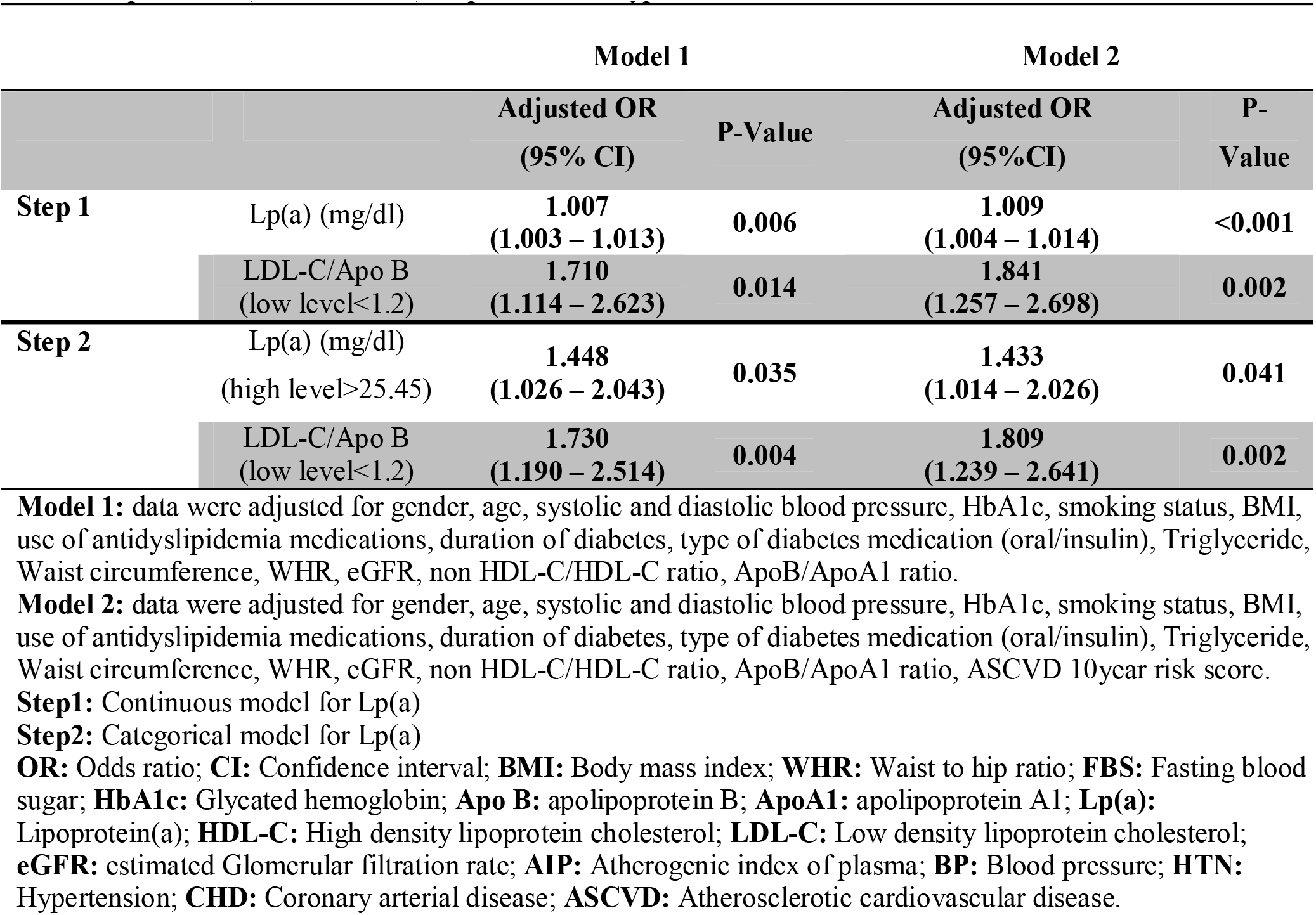
Binary logistic regression analysis determining the association between CHD status, Lp(a) (mg/dl), LDL-C/ApoB ratio (low level <1.2), in patients with type 2 diabetes

|  |  | Model 1 |  | Model 2 |  |
| --- | --- | --- | --- | --- | --- |
|  |  | Adjusted OR<br>(95% CI) | P-Value | Adjusted OR<br>(95%CI) | P-Value |
| **Step 1** | Lp(a) (mg/dl) | **1.007**<br>**(1.003 – 1.013)** | **0.006** | **1.009**<br>**(1.004 – 1.014)** | **<0.001** |
|  | LDL-C/Apo B<br>(low level<1.2) | **1.710**<br>**(1.114 – 2.623)** | **0.014** | **1.841**<br>**(1.257 – 2.698)** | **0.002** |
| **Step 2** | Lp(a) (mg/dl)<br>(high level>25.45) | **1.448**<br>**(1.026 – 2.043)** | **0.035** | **1.433**<br>**(1.014 – 2.026)** | **0.041** |
|  | LDL-C/Apo B<br>(low level<1.2) | **1.730**<br>**(1.190 – 2.514)** | **0.004** | **1.809**<br>**(1.239 – 2.641)** | **0.002** |

**Model 1:** data were adjusted for gender, age, systolic and diastolic blood pressure, HbA1c, smoking status, BMI, use of antidyslipidemia medications, duration of diabetes, type of diabetes medication (oral/insulin), Triglyceride, Waist circumference, WHR, eGFR, non HDL-C/HDL-C ratio, ApoB/ApoA1 ratio.
**Model 2:** data were adjusted for gender, age, systolic and diastolic blood pressure, HbA1c, smoking status, BMI, use of antidyslipidemia medications, duration of diabetes, type of diabetes medication (oral/insulin), Triglyceride, Waist circumference, WHR, eGFR, non HDL-C/HDL-C ratio, ApoB/ApoA1 ratio, ASCVD 10year risk score.
**Step1:** Continuous model for Lp(a)
**Step2:** Categorical model for Lp(a)
**OR:** Odds ratio; **CI:** Confidence interval; **BMI:** Body mass index; **WHR:** Waist to hip ratio; **FBS:** Fasting blood sugar; **HbA1c:** Glycated hemoglobin; **Apo B:** apolipoprotein B; **ApoA1:** apolipoprotein A1; **Lp(a):** Lipoprotein(a); **HDL-C:** High density lipoprotein cholesterol; **LDL-C:** Low density lipoprotein cholesterol; **eGFR:** estimated Glomerular filtration rate; **AIP:** Atherogenic index of plasma; **BP:** Blood pressure; **HTN:** Hypertension; **CHD:** Coronary arterial disease; **ASCVD:** Atherosclerotic cardiovascular disease.

### 3.3. AIP and CHD

The Pearson correlation analysis indicated that AIP significantly positively correlated with all CHD risk factors included: age, triglyceride, total cholesterol, LDL-C, WHR, BMI, ASCVD score, HbA1c, and negatively correlated with HDL-C (Table4). However, AIP did not have a significant correlation with LDL-C/ApoB ratio (P=0.414). Multinomial regression analysis showed it is more likely that patients have a higher risk score for CHD (borderline or moderate risk category compared to low-risk category)if they have a higher level of AIP, based on the ASCVD 10-year risk score interpretation (OR=2.236, P=0.009; OR=3.053, P<0.001; respectively), albeit patients categorized in the high-risk category compared to patients in the low-risk category have clinically higher, but not statistically significantly higher, levels of AIP (OR=6.064, P=0.053) (Table 5). Regardless, the AIP level could still not predict CHD and there was no significant difference in the AIP level between CHD positive and negative groups (P=0.803) (Figure 3).

**Table 4.** Correlation between AIP and other variables

|  | **r** | **P-Value** |
| --- | --- | --- |
| **Age** | **-0.206** | **<0.001** |
| **SBP** | 0.014 | 0.648 |
| **DBP** | **0.119** | **<0.001** |
| **Triglyceride** | **0.869** | **<0.001** |
| **Total Cholesterol** | **0.212** | **<0.001** |
| **LDL-C** | **0.067** | **0.030** |
| **HDL-C** | **-0.604** | **<0.001** |
| **non HDL-C** | **0.379** | **<0.001** |
| **non HDL-C / HDL-C** | **0.718** | **<0.001** |
| **Lp(a)** | **-0.072** | **0.021** |
| **Apo B** | **0.125** | **<0.001** |
| **Apo A1** | **-0.174** | **<0.001** |
| **Apo B / Apo A1** | **0.152** | **<0.001** |
| **LDL-C / Apo B** | -0.025 | 0.414 |
| **WHR** | **0.167** | **<0.001** |
| **BMI** | **0.123** | **<0.001** |
| **HOMA-IR** | **0.238** | **<0.001** |
| **ASCVD 10y risk score** | **0.116** | **<0.001** |
| **HbA1C** | **0.063** | **0.040** |
| **eGFR** | **0.185** | **<0.001** |
| **CRP** | 0.013 | 0.741 |

**BMI:** Body mass index; **WHR:** Waist to hip ratio; **FBS:** Fasting blood sugar; **HbA1c:** Glycated hemoglobin; **HOMA-IR:** Homeostatic Model Assessment of Insulin Resistance; **CRP:** C-reactive protein; **ApoB:** apolipoprotein B; **ApoA1:** apolipoprotein A1; **Lp(a):** Lipoprotein(a); **HDL-C:** High density lipoprotein cholesterol; **LDL-C:** Low density lipoprotein cholesterol; **eGFR:** estimated Glomerular filtration rate; **AIP:** Atherogenic index of plasma; **BP:** Blood pressure; **HTN:** Hypertension; **CHD:** Coronary arterial disease; **ASCVD:** Atherosclerotic cardiovascular disease. **r:** Pearson correlation coefficient.

**Table 5.**
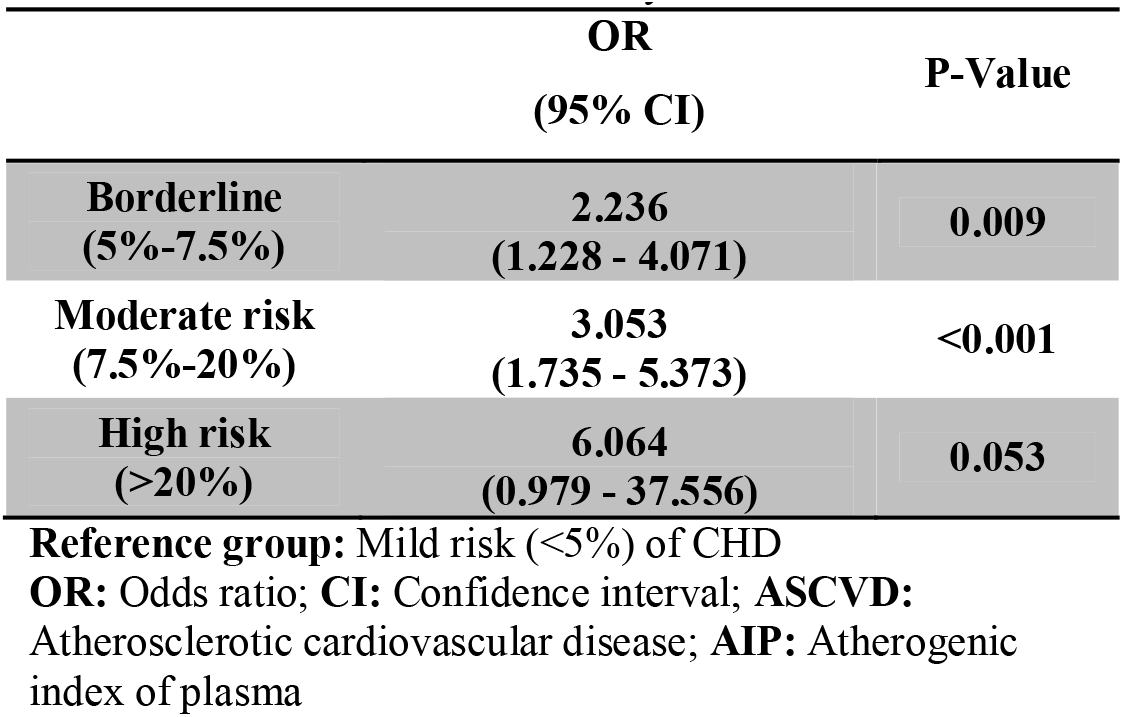
Multinomial regression analysis determining the association between ASCVD 10year risk score and AIP

|  | **OR**<br>**(95% CI)** | **P-Value** |
| --- | --- | --- |
| **Borderline**<br>**(5%-7.5%)** | **2.236**<br>**(1.228 - 4.071)** | **0.009** |
| **Moderate risk**<br>**(7.5%-20%)** | **3.053**<br>**(1.735 - 5.373)** | **<0.001** |
| **High risk**<br>**(>20%)** | **6.064**<br>**(0.979 - 37.556)** | **0.053** |

**Reference group:** Mild risk (<5%) of CHD
**OR:** Odds ratio; **CI:** Confidence interval; **ASCVD:** Atherosclerotic cardiovascular disease; **AIP:** Atherogenic index of plasma

## 4. Discussion

This case-cohort study demonstrates that low LDL-C/Apo B ratio and Lp(a) predicts CHD independent of ASCVD risk score when adjusted for multiple confounders, including lipid and glycemic indices.

### 4.1. LDL-C /ApoB ratio

Although the association between lipids and CHD has been confirmed before, the continuing question about which lipoprotein subfraction is the most atherogenic and predictive remains unresolved.

Small dense LDL-particles are highly atherogenic due to lower binding affinity for the LDL receptor, more penetration into the arterial wall, lower resistance to oxidative stress, and prolonged half-life [9, 10]. ApoB is the major structural lipoprotein in LDL-C, very low-density lipoprotein, intermediate density lipoprotein, and LP(a). An LDL-C/ApoB ratio lower than 1.2 may be used for distinguishing large buoyant LDL from small dense LDL. The incidence of CHD increases significantly in patients with small LDL-particle sizes[11, 12, 24, 25]. In the current cohort study, the results of the logistic regression analysis revealed that low LDL-C/Apo B ratio is an independent predictor of CHD. A case-control study by Tani et al. [26] showed that patients with CHD had a significantly lower level of LDL-C/ApoB ratio compared to CHD negative subjects in male patients treated with statins. Consistent with these results, another cross-sectional study by Tani et al. [25] showed that in patients with diabetes the LDL-C/ApoB ratio was lower in patients with CHD compared to those without CHD. In another case-control study, Biswas et al.[27]showed that the LDL-C/Apo B ratio was lower in the non-diabetic CHD group compared with healthy controls, but the difference was not statistically significant, though LDL-particle size had an odds ratio of 9.54 for CHD.

Kaneva et al. [12]studied the LDL-C/ApoB ratio in 186 healthy men, of which 26.3% had a ratio ≤1.2. Other studies have shown that the prevalence of phenotype B (predominance of sd-LDL particles) is 30–35% in adult men, 5–10% in men <20 years old and in pre-menopausal women, and 15–25% in postmenopausal women[28, 29]. The current study showed that the prevalence of an LDL-C/Apo B ratio ≤1.2 was 54% in CHD negative (55.7% of males, 52.6% of females) and 66.9% in CHD positive (67.8% of males,65.3% of females) patients with T2D.

Cardiovascular disease risk prediction tools such as the ASCVD risk score, based on the presence of known CVD risk factors, could estimate the probability of cardiovascular events within a defined timeframe[30]. This score was designed to identify appropriate candidates for statin therapy to prevent cardiovascular events based on an array of cardiovascular risk factors[31]. Most recent American College of Cardiology/American Heart Association guidelines on the primary prevention of CVDrecommendsconsiderationofother risk factors, including triglycerides, hsCRP, Lp(a) and Apo B, to enhance decision-making for the initiation or intensification of statin treatment[32].

A relationship between Lp(a) and the progression of diabetic complications including CHD, nephropathy, and neuropathy was recently reported by our group[33]. In the current study Lp(a) was examined by ROC analysis to derive a predictive value for CHD using a logistic regression model after adjustment for multiple confounding factors and a 10-year ASCVD risk score. High Lp(a) levels > 25.45 mg/dl were found to be independent predictor for CHD.

Although in this study the ApoB level alone was not statistically significant for the prediction of CHD, a lower LDL-C/ApoB ratio, pragmatically as an alternative to using LDL-particle size alone,does predict CHD independent of the ASCVD risk score. By extension, the LDL-C/ApoB ratio may be clinically useful to predict CVD events and prompt earlier preventive action.

### 4.2. AIP and CHD

The outcomes of this study showed that AIP has a significant correlation with some of the CHD risk factors, including older age, higher TG, LDL-C, BMI, WHR,andA1C, as well as a low level of HDL-C. In addition, the higher level of AIP was strongly associated with a higher 10-year CVD risk score. However, the AIP in this study could not predict CHD events. These findings agree with the Niroumand et al. study results, which showed in cross-sectional study design in the Iranian general population that AIP was significantly correlated with certain CHD risk factors (e.g., high BMI, high WC, and low physical activity)[34]. It is important to note that previous research studies suggested that AIP may be recommended for the prediction of CHD in the general population [35-38].

The different results in AIP’s predicting ability of CHD and AIP’s correlation with small dense LDL-C particles in this study, compared to previous studies, maybe due to different characteristics of the study population. To the best of our knowledge, there was no study reporting the predictive value of AIP for CHD in patients with type2 diabetes. This study showed that using AIP alone is not predictive for CHD in patients with type2 diabetes. Nevertheless, an observational cohort study in China of patients with T2D undergoingPCIdemonstrated that a higher AIP was strongly associated with poorer prognosis and higher incidence of major cardiovascular and cerebrovascular adverse events [39]

The characteristics of dyslipidemia in patients with T2D due to insulin resistance are postprandial chylomicronemia, elevated triglycerides and VLDL, low HDL-c, and increased small dense LDL particles. The high levels of large triglyceride-containing very low-density lipoprotein (VLDL) molecules, participate in the production of small dense LDL due to the actions of cholesteryl ester transfer protein and hepatic lipase [40-43]. According to a review by Mechanick et al. atherogenic dyslipidemia, accompanied by insulin resistance accelerate atherosclerosis independent of LDL-c levels, and begins early in the course of Cardiometabolic-Based Chronic Disease (CMBCD) [44] [45].

## 5. Conclusion

Reduction of LDL-c level is the major goal of lipid-lowering therapies but CHD continues to occur in patients with T2D. By using LDL-c reduction as the only goal of lipid-lowering therapy in patients with T2D, the opportunities to decrease the risk of atherosclerotic CHD may be missed. So, identification of other predictive lipoproteins is needed and it may have important clinical implications. This study showed that the LDL-C/ApoB ratio and Lp(a) each, but not the atherogenic index of plasma, may be considered as indicators of CHD independent of the ASCVD risk score in patients with T2D.

## Data Availability

Data are available upon reasonable request.

## List of abbreviations

SBP: Systolic blood pressure
DBP: Diastolic blood pressure
TG: Triglyceride
SD: Standard deviation
M: Male
F: Female
DM: Diabetes Mellitus
BMI: Body mass index
FBS: Fasting blood sugar
HbA1c: Hemoglobin A1C
HDL-C: High density lipoprotein cholesterol
LDL-C: Low density lipoprotein cholesterol
eGFR: estimated Glomerular filtration rate
AIP: Atherogenic index of plasma
BP: Blood pressure
HTN: Hypertension
CHD: Coronary heartdisease
ASCVD: Atherosclerotic cardiovascular disease
ACEI: Angiotensin-converting enzyme inhibitor
ARB: Angiotensin II receptor blocker
CCB: Calcium channel blocker
HCT: Hydrochlorothiazide
WC: Waist circumference
HPLC: High performance liquid chromatography
PCI: Percutaneous coronary intervention
CABG: Coronary artery bypass graft
HRT: Hormone replacement therapy
T2D: Type 2 diabetes
MS: Metabolic syndrome
WHR: Waist to hip ratio
2hpp: Two hours post prandial blood sugar
MD: Mean difference
HOMA-IR: Homeostatic Model Assessment of Insulin Resistance
CRP: C-reactive protein
ApoB: apolipoprotein B
ApoA1: apolipoprotein A1
Lp(a): Lipoprotein(a)
r: Pearson correlation coefficient
OR: Odds ratio
CI: Confidence interval

## Acknowledgments

Authors would like to appreciate the support and constructive comments of methodology research development office, ImamHospital complex, Tehran, Iran

## Licence statement

I, the Submitting Author has the right to grant and does grant on behalf of all authors of the Work (as defined in the below author licence), an exclusive licence and/or a non-exclusive licence for contributions from authors who are: i) UK Crown employees; ii) where BMJ has agreed a CC-BY licence shall apply, and/or iii) in accordance with the terms applicable for US Federal Government officers or employees acting as part of their official duties; on a worldwide, perpetual, irrevocable, royalty-free basis to BMJ Publishing Group Ltd (“BMJ”) its licensees and where the relevant Journal is co-owned by BMJ to the co-owners of the Journal, to publish the Work in Heart and any other BMJ products and to exploit all rights, as set out in our licence.

## Ethical approval

All procedures performed in studies involving human participants were in accordance with the ethical standards of the institutional and/or national research committee and with the 1964 Helsinki declaration and its later amendments or comparable ethical standards.

## Informed consent

Informed consent was obtained from all individual participants included in the study.

